# Early diagnosis of coeliac disease in the Preventive Youth Health Care Centres in the Netherlands: study protocol of a case-finding study (GLUTENSCREEN)

**DOI:** 10.1101/2021.04.24.21254842

**Authors:** Caroline R. Meijer, M. Elske van den Akker, Leti van Bodegom, Johanna C. Escher, Nan van Geloven, Floris van Overveld, Edmond H.H.M. Rings, Lucy Smit, Martine de Vries, M. Luisa Mearin

## Abstract

**Introduction:** Coeliac disease (CD) occurs in 1% of the population, develops early in life and is severely underdiagnosed. Undiagnosed and untreated disease is associated with short- and long-term complications. Treatment with a gluten-free diet results in health improvement. The current health care approach is unable to solve the underdiagnosis of CD and timely diagnosis and treatment is only achieved by active case finding. Aim of this study is to perform a novel case-finding project to detect CD in 12 months-4 years old symptomatic children who visit the YHCCs in a well-described region in the Netherlands to show that it is feasible, cost-effective and well accepted by the population.

**Methods/analysis:** Prospective intervention cohort study. Parents of all children aged 12 months-4 years attending the Youth Health Care Centres (YHCCs) for a regular visit are asked if their child has one or more CD-related symptoms from a standardized list. If so, they will be invited to participate in the case-finding study. After informed consent, a point of care test (POC) to assess CD-specific antibodies against tissue-transglutaminase (TG2A) from a droplet of blood, is performed onsite at the YHCCs. If the POC test is positive, CD is highly suspected and the child will be referred to hospital for definitive diagnosis according to the ESPGHAN guideline.

Main outcomes: 1. incidence rate of new CD diagnoses in the study region Kennemerland in comparison to the rest of the Netherlands.

2. Feasibility and cost-effectiveness of active case-finding for CD in the YHCCs. All costs of active case finding, diagnostics and treatment of CD and the potential short and long term consequences of the disease will be calculated for the setting with and without case finding.

3. Ethical acceptability: by questionnaires on parental and health care professionals satisfaction. A statistical analysis plan (SAP) has been written and will be published on the GLUTENSCREEN website.

**Ethics and dissemination:** The Medical Ethics Committee Leiden approved this study. If we prove that active case finding in the YHCC is feasible, efficient, cost-effective and well accepted by the population, implementation is recommended.

**Trial registration number:** NL63291.058.17

**What is already known on this topic?:**

- Despite recommendation on ‘who should be tested for CD’ in guidelines, the diagnosis of CD remains severely underdiagnosed.
- Untreated CD has a considerable health burden for society.
- Studies have shown that an active case-finding strategy in adults is an effective means to improve the frequency of CD diagnosis.

**What this study hopes to add?:**

- Effectiveness and feasibility of active-case finding as secondary prevention strategy in the diagnosis of childhood CD in the primary care setting in the Netherlands
- This study will provide important information about the cost-effectiveness and acceptability of the general Dutch population concerning active case-finding

## INTRODUCTION

Coeliac Disease (CD) is an immune-mediated systemic disorder elicited by the ingestion of gluten containing cereals from the normal diet (among others wheat, rye and barley) in genetically susceptible individuals. CD is characterized by a variable combination of gluten-dependent clinical manifestations, CD specific antibodies, HLA-DQ2 or HLA-DQ8 haplotypes and enteropathy[1, 2]. CD has a frequency of at least 1% in the general population, i.e. 168,000 individuals and 33,600 children in the Netherlands[3-6]. It is the most common food intolerance in the Netherlands and therefore a significant public health problem. CD is frequently unrecognized, partially because of its variable clinical presentations and symptoms, ranging from malabsorption with chronic diarrhea, poor growth in children and weight loss, to nonspecific signs and symptoms like chronic fatigue, osteoporosis/reduced bone mineral density, iron-deficiency anaemia, anorexia, chronic abdominal pain, vomiting, flatulence, irritability, elevated liver enzymes or constipation[1, 7]. CD has a considerable health burden for society. In addition to the signs and symptoms, untreated disease is associated with long-term complications such as delayed puberty, neuropsychiatric disturbances, associated autoimmune disease, miscarriages, small-for-date-births, osteoporosis, and, rarely, malignancy[1, 8]. CD increases the overall mortality risk, reduces the quality of life and yields extensive negative economic consequences, thereby presenting a resource challenge for current and future health systems[9, 10, 11].

In 1999 our research group published that childhood CD in the Netherlands was severely underdiagnosed: for every child diagnosed with CD, there were seven who have unrecognized, and therefore untreated disease[12]. Data from the National Dutch Paediatric Surveillance Unit (DPSU) show 1107 new cases in 2010-2013 of clinically diagnosed CD in children 0-14 years[13, 14]. The percentage of children diagnosed with CD <2 years of age was 30%, and < 4 years of age was 50%. Those were also the children with the most severe clinical presentations[13, 14].

DPSU is a unique registry of the Dutch Society of Paediatrics, comprising of all Dutch paediatric practices. Under it, Dutch paediatricians are asked to report newly diagnosed cases of certain diseases (CD in our case). DPSU respondents have a 90% mean response rate. The incidence of 1.56/1000 live births in 2010-2013 does not correspond to the prevalence in the general population [13, 15]. This illustrates that the current standard health care is not able to solve the problem. Once diagnosed, the patient’s health status improves after treatment with a gluten free diet (GFD) but prevention would be more beneficial[7, 16].

Results from recent prospective studies have shown that primary prevention of CD by improving the timing of gluten introduction and/or the duration or maintenance of breast-feeding is not possible[17-21]. For this reason, early diagnosis and treatment of CD represents the only way to (secondary) prevention. There are two approaches to achieve this: mass screening and case-finding. The Medical Ethics Committee (METC-Leiden Den Haag Delft, METC-LDD) considered the current evidence insufficient to assess the balance of benefits and harms of screening for CD in asymptomatic children (mass screening),[22, 23]. Consequently, we propose an active case finding project in symptomatic children in a Youth Health Care Centres (YHCC) region in the Netherlands to achieve secondary prevention of the disease. Active case-finding refers to liberal diagnostic testing of patients with CD-associated symptoms. In the general adult population, this approach has led to the early diagnosis of a large number of patients, resulting in significantly health improvement after treatment, good compliance with the GFD and good CD related quality of life,[24, 25].

In the Netherlands, more than 95% of all children 0 months-4 years visit the YHCCs,[26]. The goal of YHC is to promote and secure the health and safety of all children 0-18 years,[27]. YHC aims at primary and secondary prevention of diseases in order to promote healthy growth and development. Secondary prevention (early diagnosis and treatment) of CD therefore fits within the goals of YHC. The validated, rapid point of care (POC) test to determine CD specific antibodies represent a reliable, cheap, and easy-to-use instrument for CD case-finding in children,[28]. Therefore, early detection of CD by case finding in the YHCCs offers a “window of opportunity” to identify CD as soon as possible preventing more severe symptoms and complications of the disease.

### Aims and hypothesis

The aim of the present study is to perform a novel case-finding project to detect CD in 12 months-to 4 years old children who visit the YHCCs in a well-described region in the Netherlands, to show that it is feasible, cost-effective and well accepted by the population. We hypothesize that GLUTENSCREEN is feasible, cost-effective and well-acceptable by the general population. To achieve this, GLUTENSCREEN will compare the results of the case-finding strategy to the outcome of current healthcare in the diagnosis of CD in children in the rest of the country.

## METHODS AND ANALYSIS

### Study design

The study is a prospective intervention cohort study. The project started the 4th of February 2019 and will end the 1^st^ of February 2023 (with interruption of 5 months due to the COVID pandemic). All parents of children aged 12 months-4 years attending scheduled visits to the YHCCs in the region Midden and Zuid Kennemerland, to be further called “Kennemerland” will be informed. At the YHCC a standardized questionnaire on CD-related symptoms will be checked (annex 1). Symptoms are reported by the parents. Weight and growth are controlled at the YHCC. If one or more CD-associated symptoms (including growth restrictions) are present, the child is eligible for the study. The CD-related symptoms (see annex 1) are based on the recommendations of CD testing (taking into account the absence of previous laboratory or other investigations, and the age of the project population) in symptomatic children and adolescents in the Guideline Coeliac Disease of the European Society for Pediatric Gastroenterology, Hepatology and Nutrition (ESPGHAN)[1].

### Control population

A national control group is based on the data reported by DPSU. Dutch paediatricians are asked by the DPSU to report newly diagnosed cases of certain diseases (CD in our case) monthly during the time of this case-finding project. The CD cases are clinically diagnosed by the pediatricians to the current standard of care. DPSU respondents have a 90% mean response rate. The cases of clinically diagnosed CD in the study region will be identified by the data of the YHCC.

### Inclusion and exclusion criteria

Inclusion criteria are: 1. 12 months to 4 years of age, 2. following a gluten containing diet, 3. one or more CD-associated symptoms (annex 1), 4. parents have a sufficient knowledge of Dutch language, 5. informed consent.

Exclusion criterium: 1. diagnosed with CD

### Recruitment and procedure

Eligible children will be identified by the YHCC administration. During 2.5 years, the parents/legal guardians (from this point on called “parents”) will receive an advance invitation from the YHCC Kennemerland with information about the study. During the regularly scheduled visit at the YHCC, the nurse or the doctor will check the symptoms list (annex 1); if one or more CD-associated symptoms are present, the nurse/doctor will give the parents the information letter and informed consent form and, after informed consent is given, she/he will make a new appointment to perform the POC. The POC test for TG2A will be performed. The symptoms list and informed consent form will be stored in a separate file in the child’s electronic record.

### Intervention

After informed consent a validated POC test to determine CD specific antibodies (TG2A, Celiac Quick Test; BioHit Oyj, Finland) which is also suitable for Immunoglobulin A (IgA)-deficient patients will be performed. It requires 1 drop of fresh blood, obtained by finger-prick. The result (positive/negative) should be interpreted after 10 minutes. If the result is negative (no TG2A) the child is considered to not have CD and the procedure is finished for this child. If the POC test is positive, the child will be referred to the paediatric-gastroenterologist for further investigation for CD diagnosis at the Outpatient Clinic of the Department of Paediatric-Gastroenterology of the Leiden University Medical Center (LUMC) in the following 3 weeks. In the LUMC, CD will be diagnosed according to the ESPGHAN guidelines,[1, 2]. A second visit (face-to-face or by telephone, depending on parental preference) will be scheduled 14 days later to discuss results. There are 3 possible outcomes:

1. CD ruled out: No further follow-up is needed.
2. CD likely, but unproven; diagnostic duodenal biopsies are advised.
3. CD is diagnosed. The patient/parents will be counselled on treatment and follow-up.

If an endoscopy to obtain duodenal biopsies under general anaesthesia is advised, the parents will receive written information on the procedure, as all other parents do in the outpatient clinic when this procedure is advised. Parents have to give oral informed consent for this procedure, and this will be noted in the patient’s medical record. The procedure will be carried out per usual LUMC regulations. Biopsies will only be performed when medically indicated for the child and not just for purpose of scientific research.

### Training and protocol adherence

To perform the POC test, the YHCC healthcare professionals followed a training provided by the employees of BioHit and according to the manufacturer’s instructions. To prevent protocol drifting they receive monthly supervision by a senior clinical physician. All POC test results are photographed and stored in the electronic patient’s file. Monthly, the researchers and the senior clinical physician of the YHCC evaluate the organization, procedure and results.

### Outcome measures

The main study outcomes are:

1. The incidence rate of new CD diagnoses in the study region Kennemerland in comparison to the rest of the Netherlands.
2. Cost-effectiveness of active case-finding of CD in the YHCCs compared to standard care.
3. Ethical acceptability: by questionnaires on parental satisfaction and health care professionals.

### Data collection

The result of the POC test will be noticed in the medical file as well as the diagnosis after further investigation. Diagnostic tools and consultations after a positive POC test will be noticed in a database and in the medical file of the child.

Parents of children who visit the YHCC and/or participate in GLUTENSCREEN, will be asked to fill in standardized questionnaires on their opinion regarding the actual case-finding and on mass screening for CD. We will ask the opinion of 1) Parents of asymptomatic children, (by definition excluded for participation in case-finding); 2) Parents who decline participation in the study; 3) Parents participating in the case-finding and 4) Parents of children with suspected CD by the case-finding procedure who will be referred to the hospital for definitive CD diagnosis. Also the health care professionals in the YHCCs with various tasks within GLUTENSCREEN will also be asked to give their opinion about the case-finding.

Costs of active case-finding, diagnostics and treatment of CD and the will be compared to the costs of diagnostics and treatment of standard care. The costs of active case-finding are the costs of discussing the symptoms list, measurement of TG2A by POC test and the diagnostic costs after a positive test (repeated TG2A measurement, endomysium antibodies (EMA), human leucocyte antigen (HLA)-typing, biopsy, paediatric consultation etc.). These costs will be measured in the prospective intervention cohort study. Cost of measurement of TG2A levels include time needed from YHC professionals and cost of test equipment and materials. Resource use after a positive test will be measured by means of a case record form. Information on diagnostic procedures of clinically diagnosed CD will be collected by the DPSU and the Dutch Coeliac Society (NCV), supplemented with parent questionnaires on healthcare use outside the hospital. Health care use will be valued according to the Dutch guideline for costing research[29].

In addition, an estimate for the costs of long-term consequences of undiagnosed CD as delayed puberty, neuropsychiatric disturbances, dental enamel hypoplasia, associated autoimmune diseases, miscarriages, small for date-births, osteoporosis, and (rarely) malignancy will be made based on literature. Together with the comparison of the cost of diagnosis and treatment of CD between a situation with and without case finding, this will give an estimate for the cost-effectiveness of active case-finding compared to standard care for a lifetime horizon.

### Withdrawal

Subjects can leave the study at any time for any reason if they wish to do so without any consequences. The investigator can decide to withdraw a subject from the study for urgent medical reasons. The parents of children who withdraw are asked to fill in the questionnaire on acceptability.

### Sample size

We assume that in the Dutch population outside the case-finding project, the incidence of children 1-4 years old with a diagnosis of CD equals 0.62/1000 children years. With 2.5 years inclusion period, we expected 5434 children taking the POC test would give high power (about 95%) to detect an at least two times higher incidence rate in the study region (alpha 5%). We expected 60% of the children to be symptomatic, and 60% participation of those symptomatic children in the POC testing, so 15,100 children would need to be requested for participation, in order to obtain 5434 children available for case-finding using a rapid POC test. Since the population in the YHCCs in the Kennemerland region is approximately 12,000 children/year with additional 4,000 added per year and 2.5 years of study duration was considered sufficient to achieve sufficient sample size. When in March 2020 the study had to be interrupted for 5 months due to the COVID pandemic, the sample size calculation was re-evaluated based on the results up to that moment, including the number of cases found in the study region in the first year of the study. Based on this evaluation, it was decided that the original inclusion period of 2.5 years could be retained.

### Statistical analyses

For the primary analysis, the incidence rate in the case-finding population will be calculated along with a 95% confidence interval and will be compared with the incidence rate in the Netherlands, obtained from the DPSU, in the same period assuming the latter has no sampling variability (so using the incidence rate in the rest of the Netherlands as a fixed reference value).

All costs of active case finding, diagnostics and treatment of CD and the potential short-term consequences of the disease will be calculated for the setting with and for the cost-effectiveness without active case finding. Healthcare use will be valued according to the Dutch guideline for costing research. For the acceptability descriptive and univariate logistic regression analyses will be performed comparing the answers from the different groups. Also, univariable logistic regression analysis of negative feelings and POC-result in relation to acceptability will also be done.

### Ethics approval

The study is approved by the Medical Ethics Committee of the Leiden University Medical Centre. All study data will be handled confidentially and coded with a unique study number. Only the research team will have access to the data. A data management plan is available.

## DISCUSSION

Several studies have shown that an active case-finding strategy in the primary care setting is an effective means to improve the (early) diagnostic rate of CD and to achieve secondary prevention[24, 25].

National guidelines on the diagnosis and treatment of CD published in 2008 recommend testing for CD in patients with a wide spectrum of intestinal and extra intestinal manifestations, in asymptomatic family members of CD cases and in groups with related conditions. This approach, together with the availability of reliable CD antibody tests, have led to a rise in the incidence of diagnosed CD in Dutch children from 1.21/1000 live births in 2000 to 1.56/1000 live births in 2010-2013. Nevertheless, the increased incidence rate does not closely correspond to its frequency in the general population. In the Generation-R project, a population based prospective cohort study, the prevalence of CD at 6 years of age was 1.5%. Due to the shift in CD presenting symptoms towards a milder form, the delay from first symptoms to CD diagnosis has been reported to be unacceptably long, at between 5–10 years for many persons and so the need for earlier diagnosis has been advocated. Early diagnosis is expected to reduce serious clinical CD. Data from the DPSU shows that 50% of the 1107 new cases of clinically diagnosed CD in children aged 0-14 years between January 2010 and December 2013 were < 4 years. These young children had the most severe symptoms of CD, including chronic diarrhoea and weight loss (71.0%) or wasting/failure to thrive (65.9%),[13, 14]. Therefore, with active case finding we aim to prevent the most serious manifestations of childhood CD.

Our study has several strengths: first, to the best of our knowledge, this is the first initiative for active case finding in the general population in the Netherlands. Since the majority of the children aged 1-4 years visit the YHCC, the study will provide insight into the incidence of childhood CD in symptomatic children in the Netherlands. Second, the actual health costs of the diagnosis of childhood CD and the cost-effectiveness of active case-finding in the Netherlands have never been prospectively investigated. Third, this study will provide important information about the acceptability of the general Dutch population concerning active case finding and in addition about the willingness of parents of asymptomatic children to participate in a mass screenings project on CD.

It would also have been interesting to explore the possibility of HLA determination at the YHCCs. Since more than 95% of CD patients carry these HLA haplotypes, their presence is valuable in identifying the population that may develop CD. In the Netherlands, about 40% of the general population is HLA DQ2 or DQ8 positive and the presence of these haplotypes is thus not discriminative for the disease. On the other hand, repeated CD testing will be unnecessary in HLA-DQ2/DQ8 negative individuals. However, HLA-DQ typing currently present important drawbacks for it to be used outside the hospital. There are no rapid tests since DNA preparation takes time. Material for DNA extraction can be obtained from whole blood (minimum quantity 4-5 ml) or from other cells, such as cheek mucosa. Venepunctures are not feasible at YHCCs. Obtaining cheek cells by smoothly brushing the buccal mucosa is a possibility, but the necessary mechanisms to store and transport the material poses logistical and economic challenges. The costs of transport, DNA extraction, HLA-typing and distribution of tests results are likely to increase the costs of the active case-finding.

## Data Availability

Data Management Plan for this study is available.

## Authors’ contributions

MLM designed and supervised the trial. MLM wrote the grant proposals and helped in designing the trial. CM drafted this paper, which was edited and modified by MLM. LS is responsible for supervision of the health care professionals at the YHCCs. The health care professionals were trained according to the manufacturer’s protocol by employees of Biohit. All authors read and approved the final manuscript.

## Ethics and dissemination

The study is approved by the Medical Ethics Committee of the Leiden University Medical Centre.

## Trial registration number

NL63291.058.17 www.glutenscreen.nl

## ABBREVIATIONS

CD: Celiac disease
CME-LUMC: Medical Ethics Committee of the Leiden University Medical Centre
DPSU: Dutch Pediatric Surveillance Unit. In Dutch: Nederlands Signalerings Centrum Kindergeneeskunde, NSCK
EMA: Endomysium antibodies
ESPGHAN: European Society for Pediatric Gastroenterology Hepatology and Nutrition
GFD: gluten free diet
HLA: human leucocyte antigen
IgA: immunoglobulin A
LUMC: Leiden University Medical Centre
METC-LDD: Medical Ethics Committee- Leiden Den Haag Delft
NCV: Dutch Coeliac Society
POC: point of contact
TG2A: Anti-tissue transglutaminase antibodies
YHCC: Youth Health Care Centres

